# Prevalence and factors associated with effective helmet use among motorcyclists in Mysuru city of Southern India

**DOI:** 10.1101/2020.05.05.20091538

**Authors:** Naveen Kikkeri Hanumantha Setty, Gautham Melur Sukumar, Sumanth Mallikarjun Majgi, Akhil Dhanesh Goel, Prem Prakash Sharma, Manasa Brahmanandam Anand

**Author notes:** Corresponding Author: Dr. Naveen Kikkeri Hanumantha Setty Assistant Professor, Department of Community Medicine and Family Medicine, All India Institute of Medical Sciences (AIIMS), Basni phase 2, Jodhpur-342005, Rajasthan, India.

## Abstract

**Background:** Helmet use reduces the risk and severity of head injury and death due to road traffic crash among motorcyclists. The protective efficacy of different types of helmets varies. Wearing firmly fastened full face helmet termed as effective helmet use provides greatest protection. This study estimates the prevalence and factors associated with effective helmet use among motorcyclists in Mysuru, a tier II city in Southern India.

**Methodology:** Cross sectional road side observational study of 3499 motorcyclists (2134 motorcycle riders and 1365 pillion riders) at four traffic intersections was done followed by interview of random sample of 129 of the above riders. Effective helmet use proportion and effective helmet use per 100 person-minutes of observation was calculated. Multivariate logistic regression analysis was done to identify factors associated with effective helmet use.

**Results:** Prevalence of effective helmet use was 28 per 100 riders and 19.5 per 100 person-minutes of observation in traffic intersections. Specific prevalence rates was higher in riders (34.5%), female riders (51.3%), male pillion riders (30.5%) . Riders commuting for work and school and those ever stopped by the police in the past 3 months had significantly higher odds of effective helmet use.

**Conclusion:** The effective helmet use among the motorcyclists in Mysore is very low. Strict enforcement and frequent checks by the police are necessary to increase the effective helmet use.

## INTRODUCTION

Road traffic injuries are currently estimated to be the eighth leading cause of death across all age groups globally, and are predicted to become the seventh leading cause of death by 2030. [1,2] Worldwide, road traffic crashes contribute to nearly 1.35 million deaths and 50 million non-fatal injuries every year. [1,2] One fourth of road traffic deaths are among motorcyclists. [2] In high-income countries motorcycle deaths typically comprise about 12% of overall traffic deaths, in middle-income countries this more than doubles to 26% and this proportion is 34% in South-East Asian Region.[2] Two wheeler rider deaths comprise 34-71% of all accident deaths in India.[3]

Motorcycles form a high proportion of vehicle fleets in many low- and middle income countries.[1,4] In India, of the 253 million vehicles registered during 2017, 73.9% are two wheelers. [5]

Head injuries account for 88% of death among motorcyclists in low-and middle-income countries.[4] Wearing a helmet reduces the risk of head injuries by 69% and possibilities of death by 42%. However, a large proportion of motorcyclists suffer head injuries in road crash in spite of wearing helmets. There are different types of helmets and their effectiveness in preventing head injury varies. Head injury is more severe among those who wear nonstandard helmet than those who wear standard helmet. [7] Evidence indicates that traumatic brain injury and impact during road crash decreases in standard as well as full-face helmet users. [3,8–10] Full-face helmet provides facial protection in addition to head protection. [4] Also, risk of head and brain injury is high among motorcyclists with loosely fastened helmets compared to those with firmly fastened helmets. [7] Thus, using standard, full-face and properly strapped helmet termed as effective helmet use is key to reducing injuries and deaths to greatest extent in motorcycle crash. With this background we assessed prevalence and factors associated with effective helmet use among the motorcyclists in Mysuru, a tier II city in Southern India.

## METHODS

This cross-sectional study involved observation of motorcyclists and road side interviews of a random sample of the observed motorcyclists in four traffic intersections in the city of Mysuru, Karnataka, India. The four sites were identified in consultation with the traffic police and selected based on highest average traffic volume, safety and feasibility of location for observation/interview.

Date collection was done by three trained independent observers for a period of one week at each of the selected sites during August 2016. The three observers took position on the side of the road close to a traffic signal. The first and second observers recorded observations for motorcycle rider and pillion rider respectively, by observing all motorized two-wheelers moving in one direction, continuously for 90 minutes from 4:30 PM to 6:00 PM (peak hour). Any helmet use and helmet usage pattern (use of standard or non-standard helmet, full-face helmet or open-face and whether the helmet was firmly fastened or not) and gender was noted. If more than one motorcycle was passing at the same time, data was captured for the motorcycle that is closest to the side of the road. Validated data collection formats developed by Wadhwaniya et.al. [11] was adapted for recording the observations.

The third observer randomly stopped the motorcycle that is closest to the side of the road passing away from the first two observers. The purpose of the study was explained and motor cycle riders were included in the study after obtaining the informed verbal consent. A validated questionnaire developed by Wadhwaniya et.al was adapted for the interview. [11] The investigator administered a set of questions consisting of age and educational status of the rider, ownership of the motorcycle, factors important while purchasing the helmet, cost of the helmet, place of purchase of the helmet, purpose of the trip, do you always wear helmet, reasons for wearing or not wearing the helmet always and in the past 3 months have they ever been stopped by the police to check helmet use. Data collectors were trained both in the class room and in the field.

The following definitions were used in the study

Standard helmet: which is either a full face or open face helmet

Non-standard helmet: refers to helmets that were designed for another purpose (Horse riding helmet, construction helmet), half-coverage helmet, which is not open-face or full-face helmet.

Proper helmet use constitutes wearing standard helmet (Open/full-face) and firmly strapped.

Effective helmet use constitutes wearing standard, full-face and firmly strapped helmet.

The project proposal was submitted and approval was obtained from the Institution Ethics Review Board (IERB) of Mysore Medical College and Research Institute (MMC & RI), Mysuru in the state of Karnataka, India. Permission was also obtained from Mysuru City traffic police.

Data was entered into Microsoft excel sheet and analysed using SPSS version 23.0. Over a period of four weeks 2134 motorcycle riders and 1365 pillion riders were observed. Among the motorcycle riders observed 129 were interviewed. Prevalence of any helmet use, standard, full-face, proper and effective helmet use per 100 motorcyclists was calculated. Specific prevalence rates provided for rider, pillion rider, male and female. Person-minutes of observation was calculated as number of observers X minutes of observation X number of days of observation. Single observer observed the motorcycle riders for 90 minutes every day for 28 days giving rise to 2520 person minutes of observation. This served as denominator for calculating the helmet use for riders and pillion riders. While calculating the same indicators for total motorcyclists (riders+pillion riders) the denominator used was 5040 person minutes of observation (2 observers × 90 minutes × 28 days). Similarly, violation of helmet use was expressed per 100 person-minutes of observation. Z test for difference between two proportions, chi-square test for categorical variables was calculated. Multivariate analysis was done using stepwise forward (likelihood ratio) binary logistic regression method with level of significance set at 0.05.

## RESULTS

### Prevalence and patterns of helmet use

Among the total motorcyclists (n=3499) only 28.1% were effective helmet users. Effective helmet use was significantly higher among motorcycle riders in comparison to pillion riders (p<0.001). Also, significantly higher proportion of riders were proper (p<0.001), full-face (p<0.001), standard (p<0.001) and any helmet (p<0.001) users as compared to the pillion riders (Table 1 and Figure 1).

**Table 1.** Distribution of riders and pillion riders based on helmet use

|  | **Riders**<br>**(n=2134)** | **Pillion riders**<br>**(n=1365)** | **Total**<br>**(n=3499)** | **p value\*** |
| --- | --- | --- | --- | --- |
|  | No. (%) | No. (%) | No. (%) |  |
|  | Use per 100 riders or pillion riders observed |  |  |  |
| Helmet use | 1961 (91.9) | 915 (67.0) | 2876 (82.2) | <0.001 |
| Standard Helmet use | 1278 (59.9) | 549 (40.2) | 1827 (52.2) | <0.001 |
| Full-face helmet use | 1106 (51.8) | 423 (31.0) | 1529 (43.7) | <0.001 |
| Proper helmet use | 880 (41.2) | 354 (25.9) | 1234 (35.3) | <0.001 |
| Effective helmet use | 737 (34.5) | 247 (18.1) | 984 (28.1) | <0.001 |

Proper helmet use= those using standard helmet and firmly strapped
Effective helmet use = those using standard, full-face helmet and firmly strapped
\*Z test for difference between proportions

**Fig. 1.**
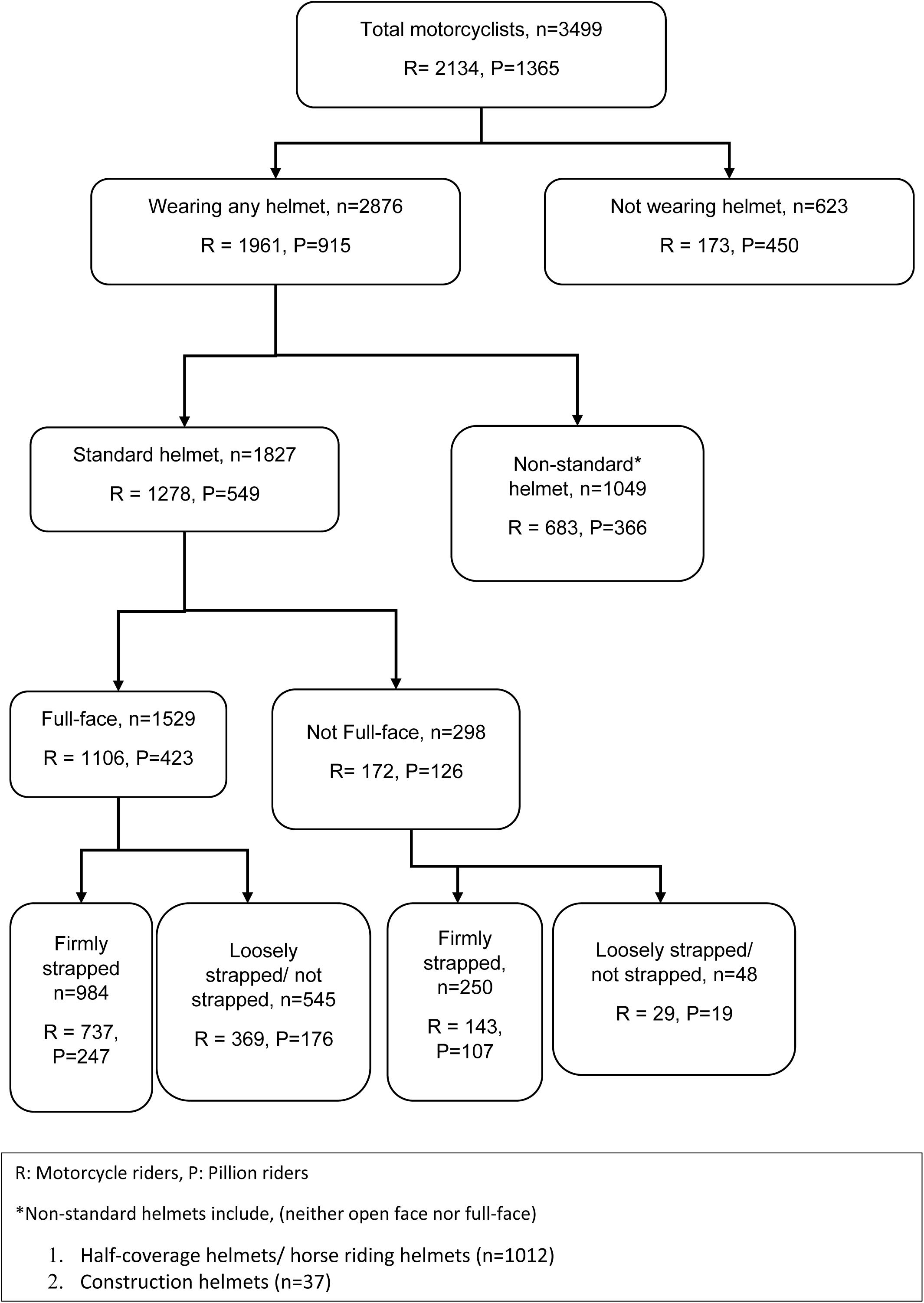
Helmet usage pattern among motorcyclists

The effective helmet use among all the motorcyclists was 19.5 per 100 person-minutes of observation and among riders and pillion riders it was 29.2 and 9.8 per 100 person-minutes of observation respectively (Table 2).

**Table 2.** Helmet use per 100 person-minutes of observation

|  | **Riders\*** | **Pillion riders\*** | **Total#** |
| --- | --- | --- | --- |
| Helmet use | 77.8 | 36.3 | 57.1 |
| Standard Helmet use | 50.7 | 21.8 | 36.3 |
| Full-face Helmet use | 43.9 | 16.8 | 30.3 |
| Proper helmet use | 34.9 | 14.0 | 24.5 |
| Effective helmet use | 29.2 | 9.8 | 19.5 |

Use per 100 person-minutes of observation = (Number of users X100)/D
D (Denominator)= Number of data collectors X number of days X (number of minutes/day)
\*For Riders / Pillion riders=( 1X28X90)=2520 person minutes of observation

Violations of helmet use per 100 person minutes of observation was also calculated. Among all motorcyclists non-effective helmet use was 16.7/100 person minutes of observation and it was 21.5 and 12.0 per 100 person minutes of observation among riders and pillion riders respectively (Figure 2).

**Fig. 2.**
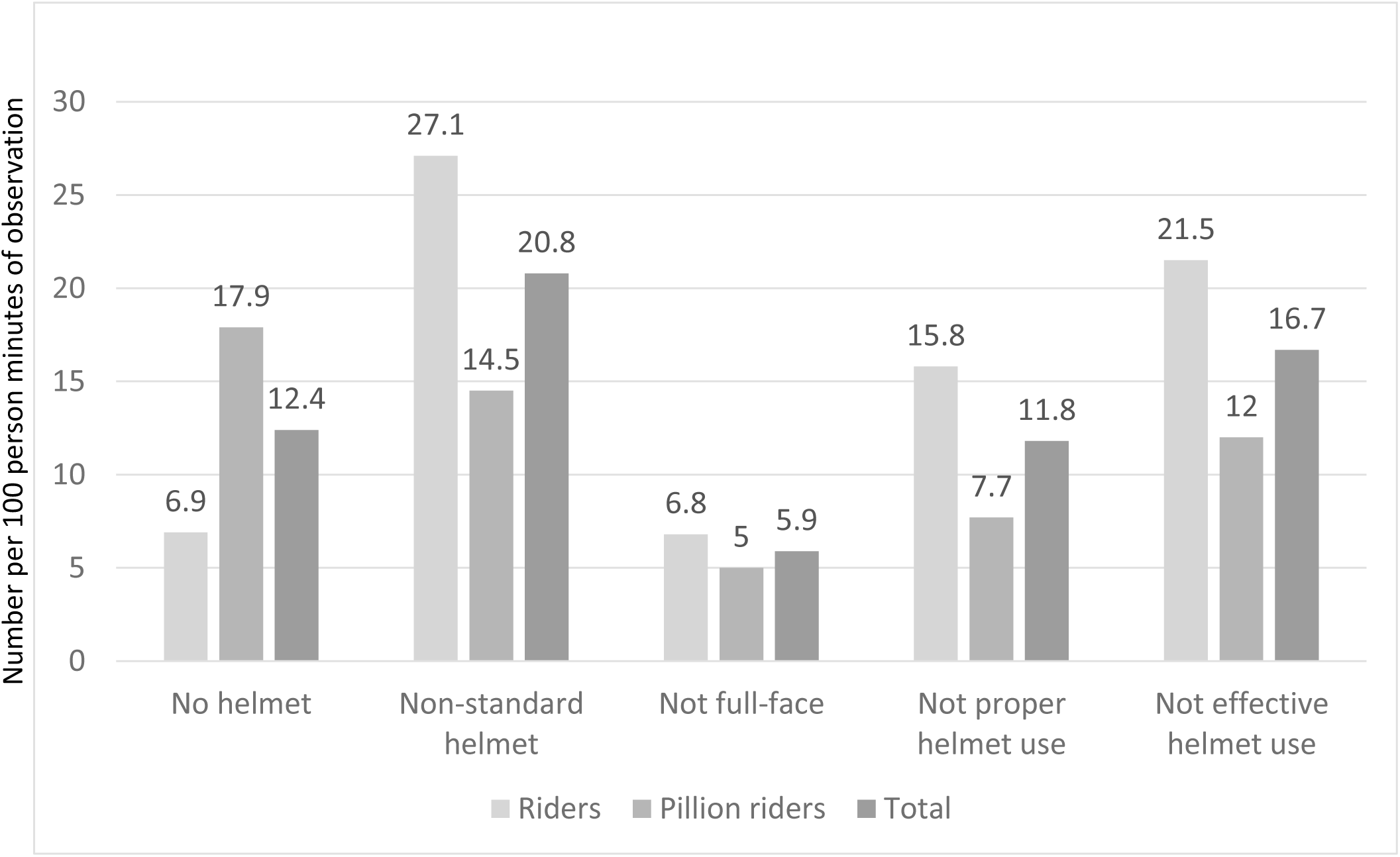
Violations of helmet use per 100 person minutes of observation

Majority of the motorcycle riders were males (n=1462, 68.5%) whereas majority of pillion riders were females (n=1008, 73.9%). All female riders were wearing any helmet while 88.2% male riders were found wearing any helmet. Significantly higher proportion of female riders were effective helmet (51.3%) users in contrast to male riders (26.8%) (p<0.001). Among the pillion riders 70.5% of female and 57.1% of males were wearing any helmet (p<0.001). However, effective helmet use was significantly higher among male pillion riders (p<0.001). On comparing the helmet usage pattern of female riders and pillion riders, significantly higher proportion of female motorcycle riders were effective helmet users as compared to female pillion riders (p<0.001). On the other hand, male motorcycle riders and pillion riders didn’t show such significant difference in their helmet use (Table 3).

**Table 3.** Distribution of riders and pillion riders based on helmet use and sex

|  | Riders (N=2134) |  | Pillion riders (N=1365) |  | P value<sup>*</sup> |  |  |  |
| --- | --- | --- | --- | --- | --- | --- | --- | --- |
|  | Male (a)<br>(n=1462) | Female (b)<br>(n=672) | Male (c)<br>(n=357) | Female (d)<br>(n=1008) | a vs b | c vs d | a vs c | b vs d |
|  | No. (%) | No. (%) | No. (%) | No. (%) |  |  |  |  |
| Helmet use | 1289 (88.2) | 672 (100.0) | 204 (57.1) | 711 (70.5) | <0.001 | <0.001 | <0.001 | <0.001 |
| Standard Helmet use | 765 (52.3) | 513 (76.3) | 177 (49.6) | 372 (36.9) | <0.001 | <0.001 | 0.352 | <0.001 |
| Full-face Helmet use | 720 (49.3) | 386 (57.4) | 167 (46.8) | 256 (25.4) | <0.001 | <0.001 | 0.401 | <0.001 |
| Proper helmet use | 418 (28.6) | 462 (68.8) | 115 (32.2) | 239 (23.7) | <0.001 | 0.002 | 0.177 | <0.001 |
| Effective helmet use | 392 (26.8) | 345 (51.3) | 109 (30.5) | 138 (13.7) | <0.001 | <0.001 | 0.159 | <0.001 |

<sup>\*</sup>Z test for difference between two proportions, vs - versus

### Characteristics of motorcycle riders interviewed

Among the 129 motorcycle riders interviewed 63 (48.8%) were aged below 30 years, 43 (33.3%) were educated upto 12th or below, 65 (50.4%) were studying or completed bachelors degree and only 21 (16.3%) were studying or completed Post graduation, 76 (58.9%) were riding motorcycle of engine capacity above 100 CC, majority owned the motorcycle (n=127, 98.4%) and 84 (65.1%) were traveling to/from work or school (Table 4). While purchasing the helmet motorcycle riders placed more importance on quality (n=97, 75.2%) and certification (n=52, 40.3%) over brand (n=27, 20.9%), style/look (n=22, 17.1%) and comfort (n=20, 15.5%). Of the 123 riders wearing helmet, certification sticker was observed in 75(58.1%) helmet, of which 73(97.3%) were authentic, 70 (56.9%) were wearing the helmet which cost rupees 500 or less (Table 4). Majority purchased their helmet from a helmet specific shop (n=88, 71.6%), few purchased from shopping mall (n=22, 17.9%) and street seller (n=10, 8.1%) and rest borrowed helmet from someone(n=3, 2.4%).

**Table 4.**
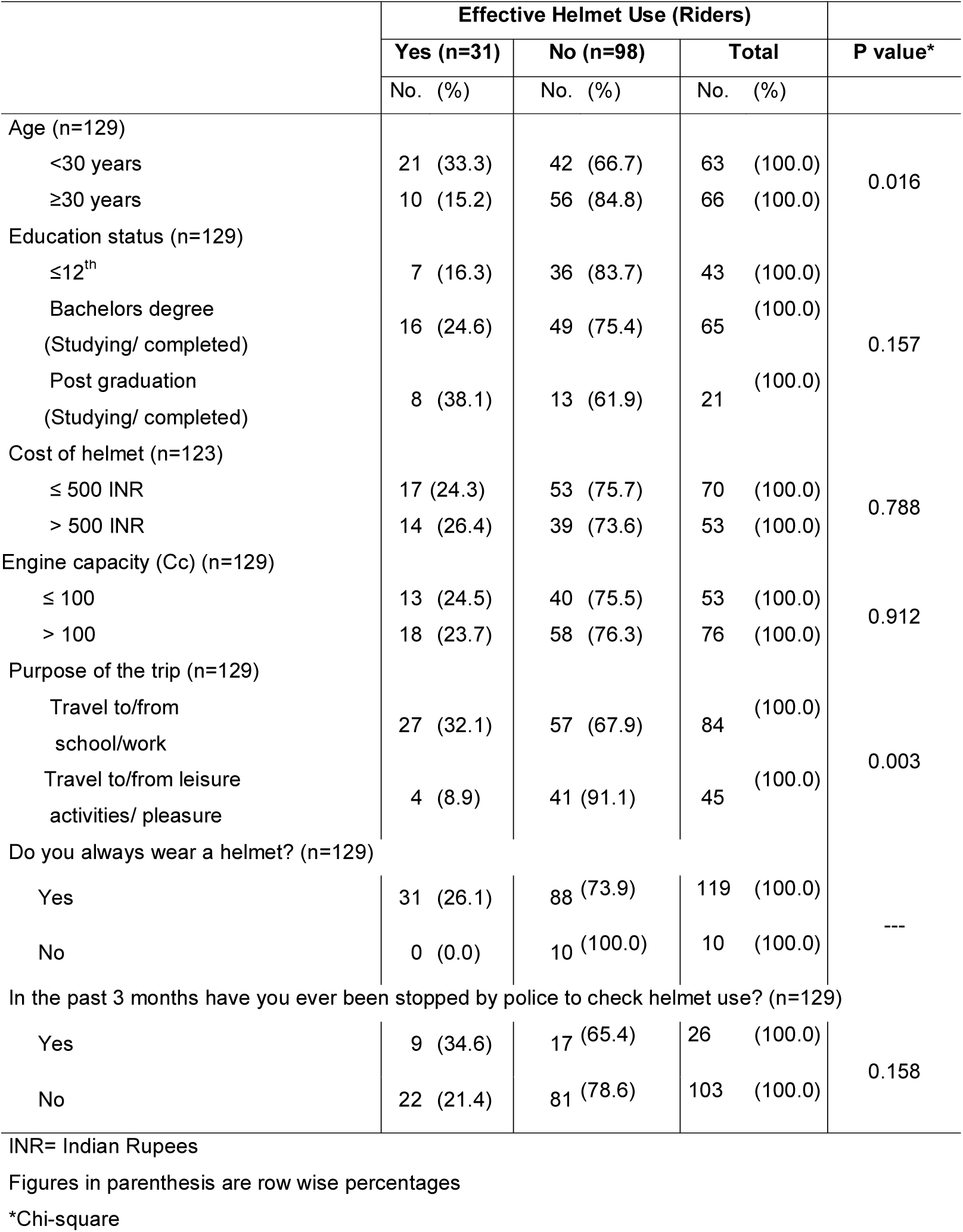
Characteristics of motorcycle riders interviewed (n=129) and effective helmet use

|  | Effective Helmet Use (Riders) |  |  | P value* |
| --- | --- | --- | --- | --- |
|  | Yes (n=31)<br>No. (%) | No (n=98)<br>No. (%) | Total<br>No. (%) |  |
| Age (n=129) |  |  |  |  |
| <30 years | 21 (33.3) | 42 (66.7) | 63 (100.0) | 0.016 |
| ≥30 years | 10 (15.2) | 56 (84.8) | 66 (100.0) |  |
| Education status (n=129) |  |  |  |  |
| ≤12<sup>th</sup> | 7 (16.3) | 36 (83.7) | 43 (100.0) | 0.157 |
| Bachelors degree<br>(Studying/ completed) | 16 (24.6) | 49 (75.4) | 65 (100.0) |  |
| Post graduation<br>(Studying/ completed) | 8 (38.1) | 13 (61.9) | 21 (100.0) |  |
| Cost of helmet (n=123) |  |  |  |  |
| ≤ 500 INR | 17 (24.3) | 53 (75.7) | 70 (100.0) | 0.788 |
| > 500 INR | 14 (26.4) | 39 (73.6) | 53 (100.0) |  |
| Engine capacity (Cc) (n=129) |  |  |  |  |
| ≤ 100 | 13 (24.5) | 40 (75.5) | 53 (100.0) | 0.912 |
| > 100 | 18 (23.7) | 58 (76.3) | 76 (100.0) |  |
| Purpose of the trip (n=129) |  |  |  |  |
| Travel to/from<br>school/work | 27 (32.1) | 57 (67.9) | 84 (100.0) | 0.003 |
| Travel to/from leisure<br>activities/ pleasure | 4 (8.9) | 41 (91.1) | 45 (100.0) |  |
| Do you always wear a helmet? (n=129) |  |  |  |  |
| Yes | 31 (26.1) | 88 (73.9) | 119 (100.0) | --- |
| No | 0 (0.0) | 10 (100.0) | 10 (100.0) |  |
| In the past 3 months have you ever been stopped by police to check helmet use? (n=129) |  |  |  |  |
| Yes | 9 (34.6) | 17 (65.4) | 26 (100.0) | 0.158 |
| No | 22 (21.4) | 81 (78.6) | 103 (100.0) |  |

INR= Indian Rupees
Figures in parenthesis are row wise percentages
\*Chi-square

Out of the 129 motorcycle riders, 119 (92.2%) said that they always wear a helmet. Most common reason for wearing the helmet always was “it can save my life” (n=103, 86.6%). Each of the rest 8(6.7%) said that they always wear helmet because police can fine them or it is required by the law. Among 10 (7.8%) respondents who answered they don’t wear the helmet always reported that most often they forget to wear (n=8, 80.0%) or they consider themselves to be a highly skilled driver (n=1,10.0%) or it is uncomfortable to wear the helmet (n=1, 10.0%). Only 26 (20.2%) motorcycle riders were ever stopped by the police to check helmet use in the past 3 months (Table 4).

### Factors affecting effective helmet use

Of the 129 motorcycle riders, 31 (24.0%) were effective helmet users. On univariate analysis age (p=0.016), purpose of the trip (p=0.003), education status (p=0.157) and being ever stopped by the police in the past 3 months (p=0.158) were factors affecting effective helmet use at p<0.20 level (Table 4).

The explanatory variables which were found significant on univariate analysis at 20% level were included in multivariate analysis. After controlling for other covariates motorcycle riders who were travelling to/from the work or school had 8.3 odds (95%CI:2.3-30.5) of wearing the helmet effectively compared to those travelling to/from leisure activities or travelling for pleasure. Those riders who were ever stopped by the police to check helmet use in the past 3 months had 4.4 odds (95%CI:1.4-14.1) of effective helmet use as compared to those who were not stopped so (Table 5).

**Table 5.** Binary logistic regression analysis – Forward step wise [LR] method

|  | **Adjusted OR [95%CI]** | **P value** |
| --- | --- | --- |
| Purpose of the trip |  |  |
| Travel to/from school/work | 8.3 [2.3 to 30.5] | 0.001 |
| Travel to/from leisure activities/ pleasure | Reference |  |
| In the past 3 months have you ever been stopped by police to check helmet use? |  |  |
| Yes | 4.4 [1.4 to 14.1] | 0.013 |
| No | Reference |  |

## DISCUSSION

The Effective helmet use in the present study among the motorcyclists (both rider and pillion rider) was 28.1%. We assessed effective helmet use as evidence indicates that chin area would see high impact in event of road crash and full face helmet provides highest protection compared to open/ half face helmet. [7,8,10] Effectiveness is better when the helmet strap is properly fastened. [7] Any deviation from this leads to lower protection and increased risk and severity of head injury. [7] There is a positive tendency of wearing any helmets as seen in 82% of the study participants. However, the use of standard (52.2%), full-face (43.7%) and proper helmet (35.3%) was low. This is comparable to other studies done in India and abroad.[12–15] With strict enforcement of helmet law there may be increase in the helmet use, however, large number of motorcyclists may wear non-standard helmet or wear them improperly. [12]

The low prevalence of effective helmet use may be due to lack of awareness of protective efficacy of different helmet types and proper fastening of strap even though motorcyclists were aware of life saving potential of helmets.[11] There are motorcyclists who habitually do not strap and another set who wear helmets only to please the traffic police and avoid the penalty. The latter group tend to remove the helmet when they move out of eyes of traffic police. Hence, they do not strap for the ease of wearing and removing the helmet with single hand while riding the motorcycle.

Over 50% of participants spent INR 500 (US $ 7.5) or less on helmet. Higher costs of standard and full-face helmets [13] prevents their use, though motorcyclists place importance on quality, certification [11] and brand of helmet while purchasing. People often tend to forget wearing helmet, however those who are engaged in routine activity like commuting to work or school are more likely to wear one [13,14] and wear it effectively. Only 20.2% of the riders were ever stopped by the police to check helmet use in the past 3 months. In Hyderabad only 2.3% of the respondents were stopped by the police. [11] Infrequent check for helmet use by traffic police and slackness in the enforcement of helmet law results in low prevalence of effective helmet use. [16] Discomfort [17] and over confidence or unrealistic optimism[18] of motorcyclists are other factors that can affect effective helmet use. Few studies have reported age and education of motorcyclists as significant factors for proper helmet use.[11,13,14] However, such association was not found in the present study.

Findings of this study indicate effective helmet use as well as other helmet use pattern was significantly lower among pillion riders. Evidence indicates lower prevalence of helmet as well as proper helmet use among pillion riders. [3,11,14] Such low prevalence among them may be due to differing perceptions of risk and compliance to law.

The effective helmet use was significantly higher among female motorcycle riders. Studies report better compliance among females.[13,14,14,19] However, as pillion riders it was significantly lower among them similar to a study from Delhi. [20] There was a significant change in the helmet use behaviour of female motorcyclists as riders and pillion riders whereas, male motorcyclists were consistent in their behaviour whether as riders or pillion riders. Even though level of risk perception is same for both sex, women were more concerned about risk of road crash [21] which justifies higher compliance. However, in Indian scenario female pillion riders are usually wife, mother, sister, daughter or aunt. They fail to adhere to helmet laws due to religious connotations, [22] gender discrimination, being less aware of the consequences, take less care of themselves or patriarchal decision making.

Novel method was tried to estimate helmet use considering human effort in denominator as number of observers and the time spent by each of them, similar to indicator assessing mosquito density per man hour of catch used in mosquito surveys. [23] Estimating helmet use per person minutes of observation can be considered as standardized indicator for such road side observation studies. It seems more meaningful when used to compare the helmet use rates across different time periods and geographical locations. With increase in helmet use it can be used to estimate violations of helmet use. Further, various violations like seat belt and helmet violations and mobile phone use while driving can be combined and compared as density of traffic violations per 100 person minutes/hours of observation. Even the CCTV footages of traffic sites can be assessed using this indicator. Use of artificial intelligence to track traffic violations can further strengthen the measurement of this indicator in a more objective manner

Towards achievement of SDGs related to road safety, target 7 was set, which aims to increase proportion of motorcycle riders correctly using properly fastened standard helmets close to 100% by 2030. [1] The enforcement level of motorcycle helmet law is 4 out of 10 points for India as per the Global status report on road safety 2018. [1] Multipronged approach is needed to reach the target. At the government level as positive note Indian Motor vehicles act was amended in August 2019, increasing fine to one thousand rupees and disqualifying licence for a period of three months to those violating the rules. [24] Strict and universal enforcement with emphasis on effective helmet use, stringent regulations prohibiting manufacture and sale of non-standard helmets and government subsidies or social marketing initiatives for standard and full-face helmets are needed. Research into development of comfortable helmets to suit local weather conditions and cost reduction and quality control by the companies. Developing alarm systems in motorcycles and helmets to remind wearing helmets and fastening strap respectively, similar to seat belt alarm in four wheelers. To implement innovative behaviour change communication strategies to public on safety potential of effective helmet use on mission mode, with special focus on pillion riders who account for 4.8-33% of two-wheeler deaths [3] and frequent monitoring by the police. Finally, addressing gender discrimination through the existing women empowerment strategies and unrealistic optimism of motorcyclists emphasizing everyone on motorcycle are at risk of head injury and death may increase effective helmet use in future.

The study is not without limitations. Observations could not be recorded for the entire day and study may not be representative of all traffic intersections in Mysuru but may approximate helmet usage pattern during evening peak traffic flow as the observation period was nearly a month. Classification of standard helmet was based solely on structure of helmets as certification sticker could not be checked for all the helmets observed. This study was done near traffic intersections which is usually monitored by the traffic police. Hence it can be overestimate of effective helmet use. The effective helmet use might be much lower in other areas which are not so monitored. Element of social desirability bias in self-reporting of helmet use cannot be ruled out.

## CONCLUSION

Out study indicates that effective helmet use in the Mysuru, a tier II city of southern India is very low and much lower among pillion riders. Disparity exists in the effective helmet use between male and female riders and pillion riders and between female riders and pillion riders. Purpose of the trip and monitoring by the police are factors associated with effective helmet use. It is recommended to strengthen enforcement, increase accessibility and affordability of low cost, high quality and comfortable standard full-face helmets along with behaviour change communication to enhance effective helmet use in India.

## KEY MESSAGES

What is already known
- Proper helmet use is low in low- and -middle income countries
- Lower prevalence of helmet use among pillion riders
- Higher prevalence of helmet use among female motorcyclists
- Strict enforcement of helmet law increasing helmet use

What this study adds
- Introduction of the term effective helmet use (standard, full-face, properly strapped helmet) and documenting its prevalence
- Lower prevalence of effective helmet use among female pillion riders
- Novel method of calculating helmet use and violations of helmet use per 100 person minutes of observation

### Declarations

#### Ethics approval and consent to participate

Institutional Ethics committee of Mysore Medical College and Research Institute, Mysuru, Karnataka, India approved the study (Letter dated: 23^rd^ July 2016)

Informed verbal consent was obtained

#### Consent for publication

Not applicable

#### Availability of data and materials

The data supporting the conclusions of this article is(are) included within the article. If the raw data is required it can be made available on reasonable request.

## Authors’ contributions

NKH, GMS, SMM and MBA conceptualized and designed the study. NKH, SMM and MBA collected the data. NKH and MBA entered the data. NKH, GMS, SMM, ADG and SPP carried out statistical analyses and interpreted the data. NKH and ADG written the first draft. GMS, SMM, ADG and SPP critically reviewed the draft for intellectual content and revised the manuscript. All the authors reviewed and approved the final draft.

## Data Availability

All data relevant to the study are included in the article or uploaded as supplementary information

## Acknowledgements

Dr Shivam Gupta, Associate Scientist, Department of International health, Johns Hopkins Bloomberg School of Public Health, Baltimore MD 21205 USA for permission to use the questionnaire and guidance and support. Mysuru city traffic police for permission, support and cooperation to conduct the study. All the motorcyclists who consented to participate in the study. Dr. Swathi. G and Dr. Varun C.R. former interns at Mysore Medical College and Research Institute, Mysuru, Karnataka, India for assisting in data collection.

